# Local ancestry modulates gene expression: shifting our understanding of genetic regulation and disease association within and across populations

**DOI:** 10.64898/2026.03.17.26348586

**Authors:** Hung-Hsin Chen, Yanwei Cai, MariaElisa Graff, Wanying Zhu, Lauren E. Petty, Heather M. Highland, Rashedah Roshani, Hannah G. Polikowsky, Anna S. Lorenz, Elizabeth Frankel, Joshua M. Landman, Mohammad Y. Anwar, Elyn E. Franson, Jeffrey Haessler, Christy L. Avery, Kristin L. Young, Lindsay Fernandez-Rhodes, Alexander P. Reiner, Ulrike Peters, Penny Gordon-Larsen, Eric R. Gamazon, Alexandre C. Pereira, Charles Kooperberg, Chad D. Huff, Susan P. Fisher-Hoch, Joseph B. McCormick, Kari E. North, Jennifer E. Below

**Affiliations:** Institute of Biomedical Sciences, Academia Sinica, Taipei, Taiwan; Division of Genetic Medicine, Vanderbilt Genetics Institute, Vanderbilt University Medical Center, Nashville, TN, USA; Public Health Sciences, Fred Hutchinson Cancer Center, Seattle, WA, USA; Department of Epidemiology, Gillings School of Global Public Health, University of North Carolina at Chapel Hill, Chapel Hill, NC, USA; Department of Epidemiology, UT Health Houston School of Public Health, Brownsville Campus, Brownsville, TX, USA; University of Washington, Seattle WA, USA; Biobehavioral Health - BBH, Penn State University, State College, PA, USA; Department of Epidemiology, University of Washington, Seattle, WA, USA; Department of Nutrition, Gillings School of Global Public Health, University of North Carolina at Chapel Hill, Chapel Hill, NC, USA; Brigham and Women’s Hospital and Harvard Medical School, Boston, MA, USA; Department of Epidemiology, The University of Texas MD Anderson Cancer Center, Houston, TX, USA

## Abstract

Genetic variants that influence transcript abundance, called expression quantitative trait loci (eQTL), are fundamental to understanding gene regulation and disease etiology. However, eQTL studies have overlooked the influence of the ancestral origin of a gene on its regulation. We implemented a new statistical framework that maps local ancestry-specific regulatory variation, revealing pervasive ancestry-specific effects on gene expression in Hispanic/Latino and African American populations. Enriched in open chromatin regions, these variants better explain genetic disease risk in these populations. The widespread heterogeneity of local ancestry-based eQTL effects offers mechanistic explanations for inconsistencies in genomic and multi-omic studies across populations. Our findings expand existing models of gene regulation and the importance of applying local genomic context in genetic studies to advance precision medicine and address health disparities.

## Introduction

Understanding the function of genetic variation is an essential step in identifying the molecular pathways through which genetic risk factors impact complex traits and diseases.(*1–3*) Mapping regulatory effects of disease-associated genetic variants has proven critical to more precise identification of prevention and treatment targets.(*4, 5*) However, despite major advances in identifying genetic variants that influence transcript abundance, called expression quantitative trait loci (eQTL), to date eQTL discovery has ignored the influence of the ancestral history of a gene on its regulation. GTEX has served as the seminal reference dataset for gene expression studies; approximately 85% of GTEX v8 participants (n=838) are self-reported European American, with only 103 and 16 participants from African American and Hispanic/Latino populations, respectively. Both of these populations have a recent history of cross-continental genetic admixture in the last 500 years.(*6*) Recombination of admixed genomes results in a varied tapestry of genomic segments, or haplotypes, derived from the ancestral populations across the chromosomes (**Figure 1A**). Recent studies in these admixed populations have demonstrated modest differences in the mapped effects of regulatory variation by global and local ancestry on total gene expression but have failed to deconvolute transcript abundance by underlying ancestry, which we hypothesized may lead to overlooked differential effects.(*7, 8*)

**Figure 1.**
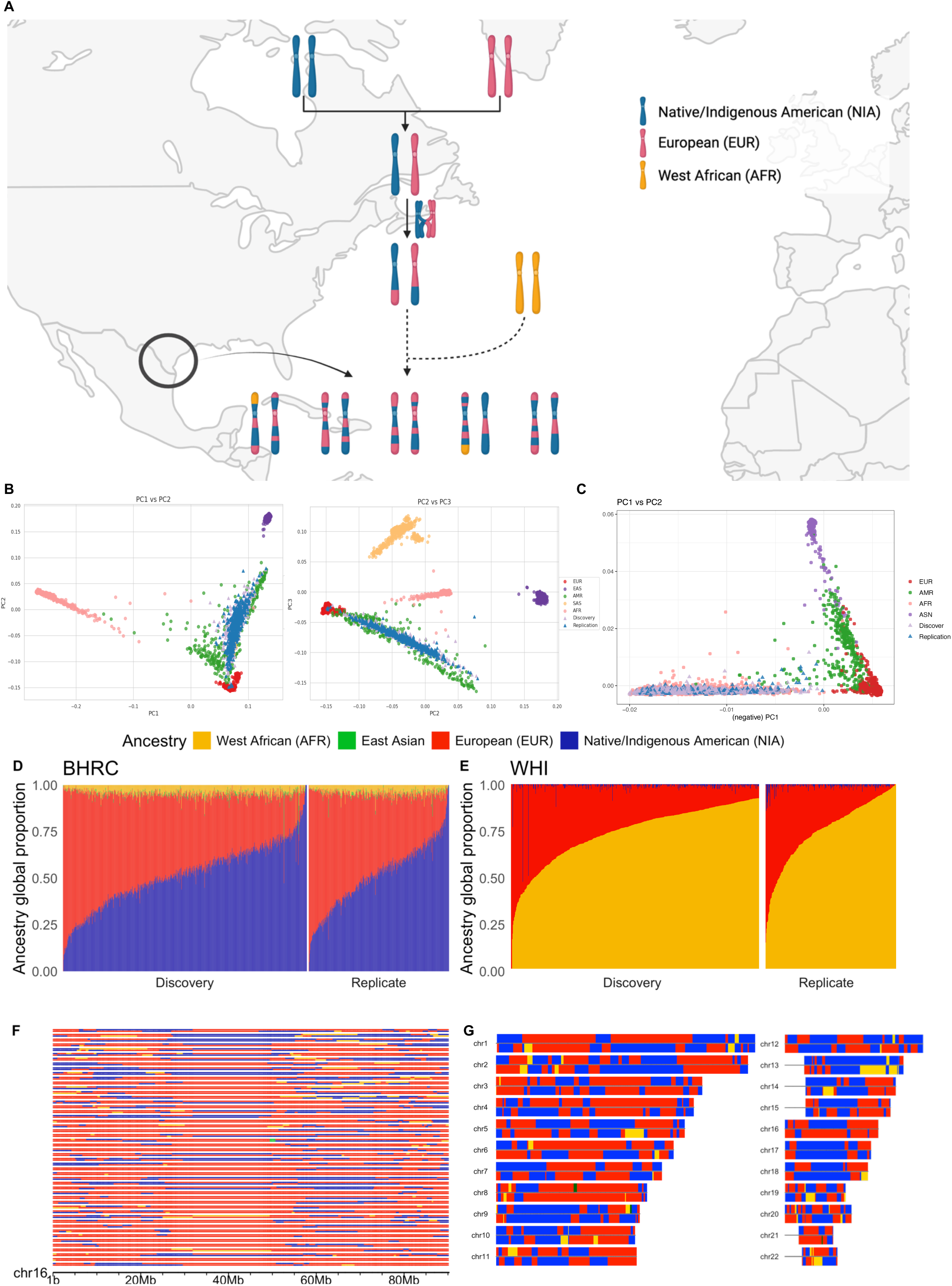
Local ancestry in the Border Health Research Cohort (BHRC) study participants. A) A simplified illustration of admixture events, resulting in the paired chromosomes for multiple individuals from an admixed population, seen at the bottom. Admixture occurs when two or more previously isolated populations merge to form a new population. Here, three ancestral populations, Native/Indigenous American (NIA), European (EUR), and West African (AFR) are represented by cyan, red, and yellow chromosomes, respectively. As generations continue, each chromosome resembles a mosaic of the initial ancestral chromosomes, as a result of recombination at meiosis. B) The projected genetic principal components analysis (PCA) plot of BHRC participants with 1000 Genomes Project participants as reference. C) The PCA plot of WHI participants in this study with all WHI participants in the TOPMed program (self-reported as EUR/AFR/Admixed American/Asian) as reference. D) Estimated global ancestry proportions of BHRC participants (autosomes) derived using ADMIXTURE and the 1000 Genomes Project reference panel and compared to global ancestry estimates from an unrelated subset of individuals from the Hispanic Community Health Study/Study of Latinos (HCHS/SOL) study population. Each vertical bar represents a single individual, and the four color-coded segments represent the three ancestry fractions. The colors depict local ancestry across each chromosome based on the 1000 Genomes Project reference panel. E) Estimated global ancestry proportions of WHI participants (autosomes) derived using ADMIXTURE. Each vertical bar represents a single individual, and the four color-coded segments represent the three ancestry fractions. F) The haplotypes of 50 randomly selected BHRC participants for chromosome 16. Each haplotype is shaded according to the local ancestry at that particular location along the chromosome, using the 1000 Genomes Project populations as the reference. G) The autosomes of a randomly selected BHRC participant, the region of determined local ancestry is limited by coverage of genotyping.

Typically, eQTL effect estimates are derived from additive allelic models that evaluate the impact of each variant irrespective of haplotype. However, allele-specific expression, which refers to the difference in expression levels between two haplotypes, has been shown to be a more powerful approach for eQTL mapping.(*9, 10*) Linking allele-specific expression and local ancestry background represents an unexplored opportunity to investigate the effects of local ancestry on gene regulation. Indeed, we show through simulations that when eQTL effects are differentiated by local ancestry, failure to model these effects greatly diminishes power to discover and predict biological effects on disease (**Supplemental Materials**). Further, because local ancestry states vary within individuals, local ancestry-based (LAB) regulatory effect estimates minimize many potential confounders.

Here, we introduce a novel statistical approach for utilizing allele-specific expression to estimate LAB eQTL effects, ancQTL. We deploy this approach to estimate LAB eQTL effects in a Hispanic/Latino population, the Border Health Research Cohort (BHRC), and an African American population, the Women’s Health Initiative (WHI), to investigate the impact of local ancestry on gene regulation (**Figure 1**).(*11*) We describe the first ancestry-specific regulatory landscape for Native/Indigenous American (NIA), African (AFR), and European (EUR) ancestries in Hispanic/Latino and African American populations. These heterogeneous regulatory effects by local ancestry context contribute to inconsistent genetic findings across admixed populations, particularly when local ancestry-specific effects are in opposite directions. Indeed, we find that LAB eQTLs improve colocalization and biological interpretation of genetic effects for a range of complex traits.

## Results

### Genome and transcriptome in BHRC

BHRC recruited participants in Brownsville, Texas, located along the United States-Mexico border, and all participants self-identify as Hispanic/Latino and 94% identify as Mexican American. We estimated their global ancestry with an average of 49.0% NIA, 45.7% EUR, and 4.6% West African (AFR) ancestry for the 962 individuals with both available genomic and transcriptomic data (**Figures 1B and D**). We then conducted bulk RNA-seq of peripheral blood samples for 645 individuals in the BHRC. Gene expression was nominally associated with the first genetic principal component (PC) (*p* < 0.05) in 3,660 genes (412 with FDR < 0.05), suggesting pervasive effects of global ancestry on RNA abundance across the genome (**Figure S2**). We then assessed differences in expression at each gene according to local ancestry at the haplotype level. We found that of the 15,262 expressed genes with continuous local genetic ancestry for both NIA and EUR in at least 20% of our participants, 838 (5.2%) were differentially expressed (FDR < 0.05) on NIA and EUR local ancestry backgrounds after adjusting for unobserved confounders measured by 60 PEER factors (**Figure 2A-B** and **Table S1**).(*12, 13*)

**Figure 2.**
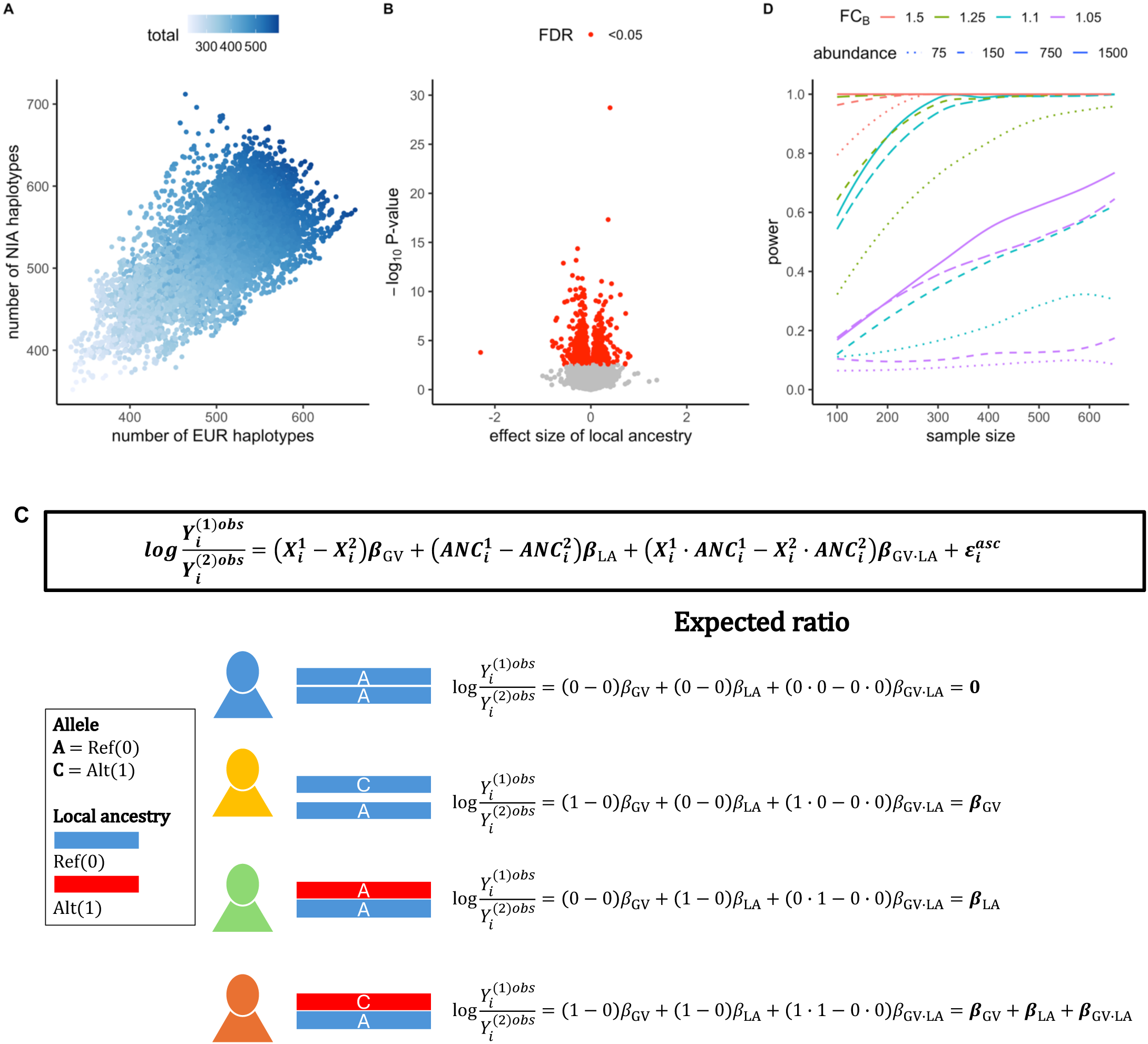
In the Border Health Research Cohort (BHRC) participants, local ancestry haplotype counts, local ancestry effect on gene expression, an illustration of our model integrating local ancestry on expression quantitative trait locus (eQTL) mapping, and power estimation for this approach. A) The scatterplot shows of number of unbroken European ancestry (EUR) (x-axis) and Native/Indigenous American (NIA) (y-axis) haplotypes for each gene; the grade of color represents the number of subjects with two unbroken haplotypes. B) A volcano plot showing the effect of local ancestry on allele-specific gene expression with red indicating false discovery rate<0.05. The x-axis shows the effect size (beta) of local ancestry on haplotype specific expression. C) Illustration of the statistical framework of integrating local ancestry in eQTL mapping, *ancQTL*. In the model, 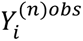 is the haplotype read count for individual *i* at haplotype n, 𝑋*_i_^n^* is the allelic dosage at the variant, 𝐿𝐴*_i_^n^* is the number of alleles at the variant in a given ancestral population, 𝜀*_i_* is the residual expression, and a vector of effect sizes 𝛽*_GV_*, 𝛽*_LA_*, 𝛽*_GV_*_∗*LA*_ representing the effect size of the genetic variant, genetic local ancestry and their interaction on the allelic expression imbalance 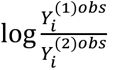. The four examples demonstrate the different scenarios of local ancestry background and alleles. D) Power of detecting the differences of local ancestry based eQTL effects (alpha<0.05) in sample sizes ranging from 100 to approximately our sample size, 650, and diverse transcript abundance (total aligned sequencing reads ranging from 75-1500). The color represents the difference of effect in the second ancestry background, with fold change ranging from 1.05 to 1.5, compared to a fixed effect in the first ancestry background (fold change=1.01). For each scenario, we conducted 200 simulations to obtain the power.

### Leveraging local ancestry with ancQTL

Under a variety of scenarios of effect size differences by local ancestry (**Figure S3**), our simulation results indicated that classical eQTL mapping results in a reduction of gene expression variance explained by 16%-52% relative to models that account for local ancestry (**Supplemental Materials**). To address this deficit, we introduce a novel statistical model, *ancQTL*, that leverages both local ancestry information and allele-specific expression to estimate ancestry-specific effects on gene expression for variants within cis-regulatory regions. Specifically, we extended the standard allelic imbalance framework (*9*) by incorporating the main effects of the genetic variant (G_V_) and genetic local ancestry (G_LA_), as well as a G_V_ x G_LA_ interaction term. Under this model, the estimated effect size of G_V_ represents the eQTL effect in the reference local ancestry background, while the effect in the alternative ancestry background is captured by the sum of the G_V_ and interaction coefficients. Furthermore, the G_V_ x G_LA_ interaction term quantifies the difference of eQTL effects between the two ancestral backgrounds. *ancQTL* exhibits high power to detect significant differences in effects by ancestral background, dependent on total aligned read count, sample size, and fold change (FC) difference between ancestry backgrounds (**Figure S4**). For example, *ancQTL* had >99% power to detect a FC difference of 0.1 (LAB eQTL FC effect of 1.01 vs. 1.10 in two ancestry backgrounds) in an average of 150 aligned reads with a sample size of 650 (**Figures 2D** and **S4**).

### LAB eQTLs in a Hispanic/Latino cohort

We conducted LAB eQTL mapping in the BHRC for the common variants (minor allele frequency [MAF] > 0.05 in both ancestry) and the 15,262 genes that passed quality control assessments (**Supplemental Materials**). In total, these analyses identified LAB eQTLs (FDR < 0.05) for 8,523 genes. These include 1,404,505 eQTLs for 6,224 genes on EUR haplotype backgrounds (EUR LAB eQTLs), and 2,178,162 eQTLs for 6,196 genes on NIA haplotype backgrounds (NIA LAB eQTLs) (**Figures S5** and **Table S2**). Of these significant eQTLs, 734,775 from 4,466 genes also had differential effect size estimates for regulation of gene expression according to local ancestry background (measured as a significant G_v_ x G_LA_ interaction effect, FDR < 0.05). Within this set, 393,138 variants exhibited ancestry-specific associations meeting the following three criteria: 1) associated with RNA abundance when appearing on haplotypes from the first ancestry group (FDR < 0.05), 2) differentially associated with RNA abundance when appearing on haplotypes from the first ancestry group versus the second ancestry group (G_V_*G_LA_ interaction FDR < 0.05), and 3) no evidence of association under the null model when appearing on the second ancestry group (*p* > 0.10) (**Figure 3A**). Among the variants with ancestry-specific associations, 101,360 LAB eQTLs mapping to 2,231 genes were specific to EUR (EUR-only) and 291,778 variants mapping to 2,968 genes were specific to NIA (NIA-only). An additional 82,026 eQTLs were identified on both ancestral backgrounds but exhibited significantly different effect sizes (G_V_*G_LA_ interaction FDR < 0.05). Of these, 44,482 eQTLs from 803 genes had the same direction of effect and 37,544 eQTLs from 1,411 genes had an opposite direction of effect. Our results, summarized in **Figures 3B-C**, demonstrate that 24.3% (734,775/3,018,633) of LAB eQTLs had a significant local ancestry-specific effect. Among the 8,523 genes evaluated, 52% had at least one such local ancestry-specific eQTL.

**Figure 3.**
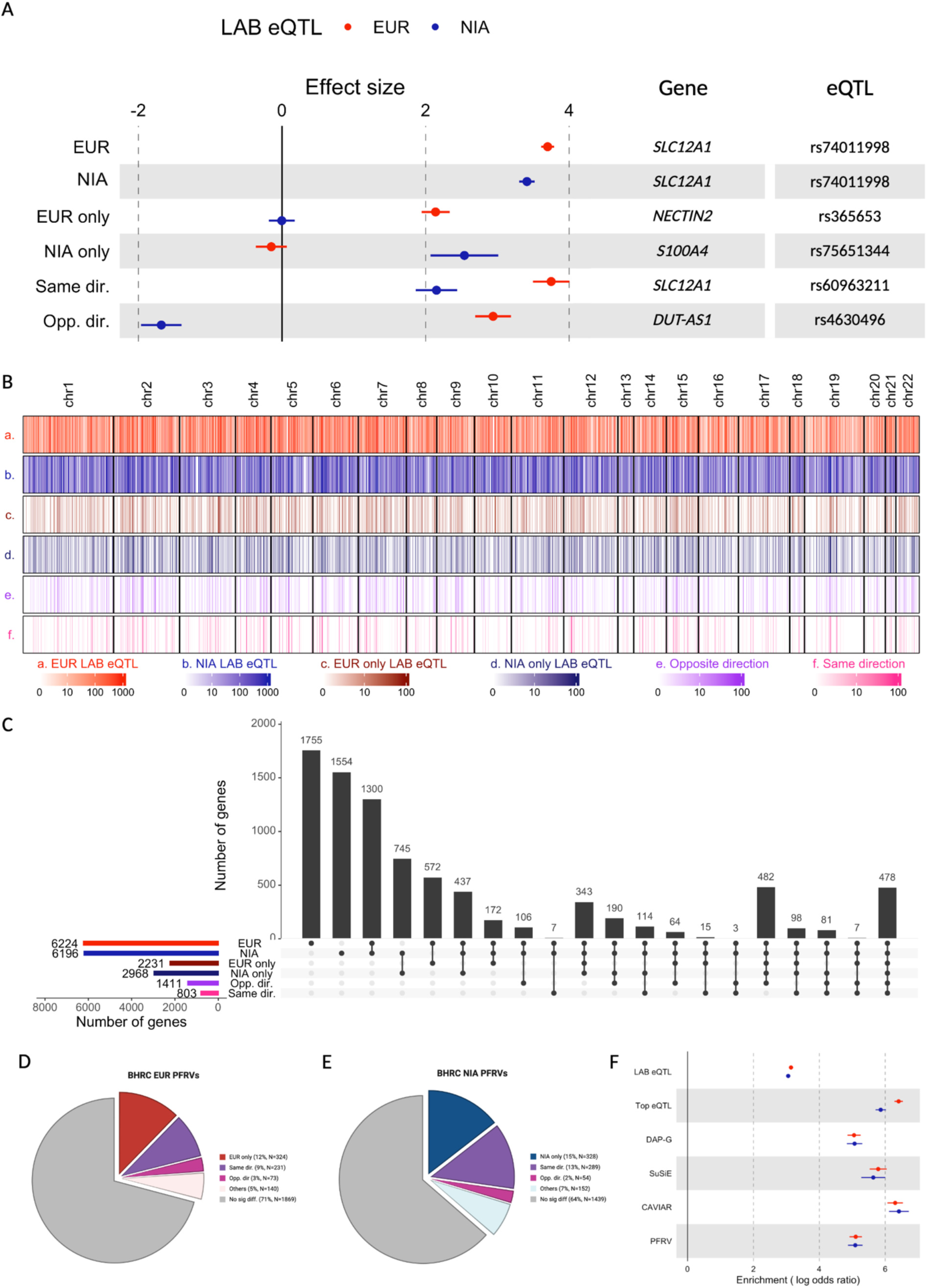
Definitions of each type of local ancestry-based expression quantitative trait locus (LAB eQTL) in the Border Health Research Cohort (BHRC) discovery set, the number of gene with different types of LAB eQTLs and their enrichment in replication. A) The plot illustrates an example of each type of local ancestry differentiated LAB eQTL, including 1) EUR and NIA LAB eQTL are defined by significant effect of the corresponding ancestry group (FDR < 0.05); 2) EUR only and NIA only LAB eQTL were defined with following criteria significant effect from the corresponding ancestry group (FDR<0.05), significat difference between two ancestry groups (G_v_ x G_LA_ FDR < 0.05), and no evidence of effect from the alternative ancestry group (*p* > 0.05); 3) significant in both ancestry groups (FDR < 0.05) but with different effect (G_v_ x G_LA_ FDR < 0.05) with same direction or opposite direction. B) The heatmap summarizes the number of identified LAB eQTL for each gene and different types of local ancestry differentiated LAB eQTLs. C) The Upset plot shows the number of genes with European ancestry (EUR) and/or Native/Indigenous American ancestry (NIA) LAB eQTLs as well as different types of local ancestry differentiated LAB eQTLs. A single gene may contain multiple types of local ancestry differentiated LAB eQTLs referring distinct loci. D) Pie chart of EUR PFRVs by ancestry-specific effect groups, as defined in panel A. E) Pie chart of NIA ancestry-specific effect groups, as defined in panel A. F) Forest plot shows the enrichment of identified LAB eQTLs in replication as well as the top and fine-mapped LAB eQTLs. The color indicates the source of local ancestry background. The enrichment is estimated with TORUS, and the bar represents the 95% confidence interval of estimation.

### Fine mapping in BHRC

The results above show that ancestry-specific eQTL effects are common in the BHRC, but they do not imply that the functional regulatory variants driving these differences behave differently according to local ancestry background. Ancestry-specific eQTLs could instead result solely from differences in haplotype structure and allele frequencies of functional regulatory variants between ancestry backgrounds. To test for ancestry-specific differences in regulatory variant function, we conducted fine-mapping to identify LAB eQTLs likely to functionally impact gene expression, hereafter termed putatively functional regulatory variants (PFRVs). We applied three fine-mapping tools, SuSiE, DAP-G, and CAVIAR, identifying PFRVs for 1,945 genes in total (defined as having one or more LAB eQTL with a posterior inclusion probability >0.8 in any fine-mapping approach, **Table S3** and **Figure S5**). These included identified 2,637 PFRVs for 1,374 genes on EUR haplotypes and 2,262 PFRVs for 961 genes from NIA haplotypes. Among these 4,011 PFRVs, 1,527 (33.5%) had ancestry-specific differences in effect, defined as significantly different effect sizes (G_v_*G_LA_ interaction FDR < 0.05). The PFRVs with ancestry-specific effects could be characterized into three common patterns, 1) functional in one ancestry with no evidence of function in the other (324 EUR-only and 328 NIA-only PFRVs), 2) functional in both ancestries with the same direction of effect but different effect sizes (465), and functional in both ancestries with opposite directions of effect (118) (**Figures 3D-E and S6**). In total, 41.9% (816/1,945) of the genes evaluated had at least one PFRV with an ancestry-specific effect (G_v_*G_LA_ interaction FDR < 0.05). Similar proportions were observed with various set intersections across the three fine-mapping tools (21.1%-27.1%).

### Replication of BHRC LAB eQTLs

We applied *ancQTL* to 317 unrelated BHRC participants to validate our identified LAB eQTLs. Of the 1,390,138 EUR LAB eQTLs in the discovery stage, 671,955 were validated in the replication set, with 48.3% of the LAB eQTLs from the discovery stage remaining significant after FDR correction. Similarly, for the NIA haplotypes, 1,037,677 out of 2,154,449 LAB eQTL were validated, with 48.2% of LAB eQTLs showing significant associations with the same direction with corresponding gene expression in our replication data following FDR correction. EUR and NIA LAB eQTLs identified in the discovery set were significantly enriched among the set of LAB eQTLs in the replication set. PFRVs exhibited substantially higher replicated enrichment ratio (odds ratio = 33.1-78.2 and 33.5-85.3 for EUR and NIA PFRVs, respectively, **Figure 3F**).

### LAB eQTLs in an African American cohort

We leveraged whole genome sequence and transcriptome data from 1,094 African American individuals from the WHI, 748 of which were included in discovery and 346 in replication. The global ancestry estimates for this cohort were 76.0% AFR, 23.1% EUR, and 1.0% NIA (**Figure 1D and 4A**). Among 8,672 genes that passed QC, we identified 871,129 LAB eQTLs in AFR haplotypes (AFR LAB eQTLs), across 4,863 genes, and 535,767 LAB eQTLs in EUR haplotypes (EUR LAB eQTLs), across 2,553 genes. The reduced number of EUR LAB eQTLs may reflect the low proportion of global ancestry of EUR in this population. Among the identified LAB eQTLs, 28,195 (for 1,088 genes) were unique to AFR haplotypes (AFR-only), and 109,175 (for 1,612 genes) were unique to EUR haplotypes (EUR-only). Additionally, 40,904 LAB eQTLs (from 1,054 genes) were observed in both ancestries but showed significantly different effect sizes, of which 19,271 LAB eQTLs (from 633 genes) showed opposite directions of effect (**Figures 4B-C** and **Table S4**). Among the 5,281 genes evaluated, 45.7% had at least one such local ancestry-specific eQTL

**Figure 4.**
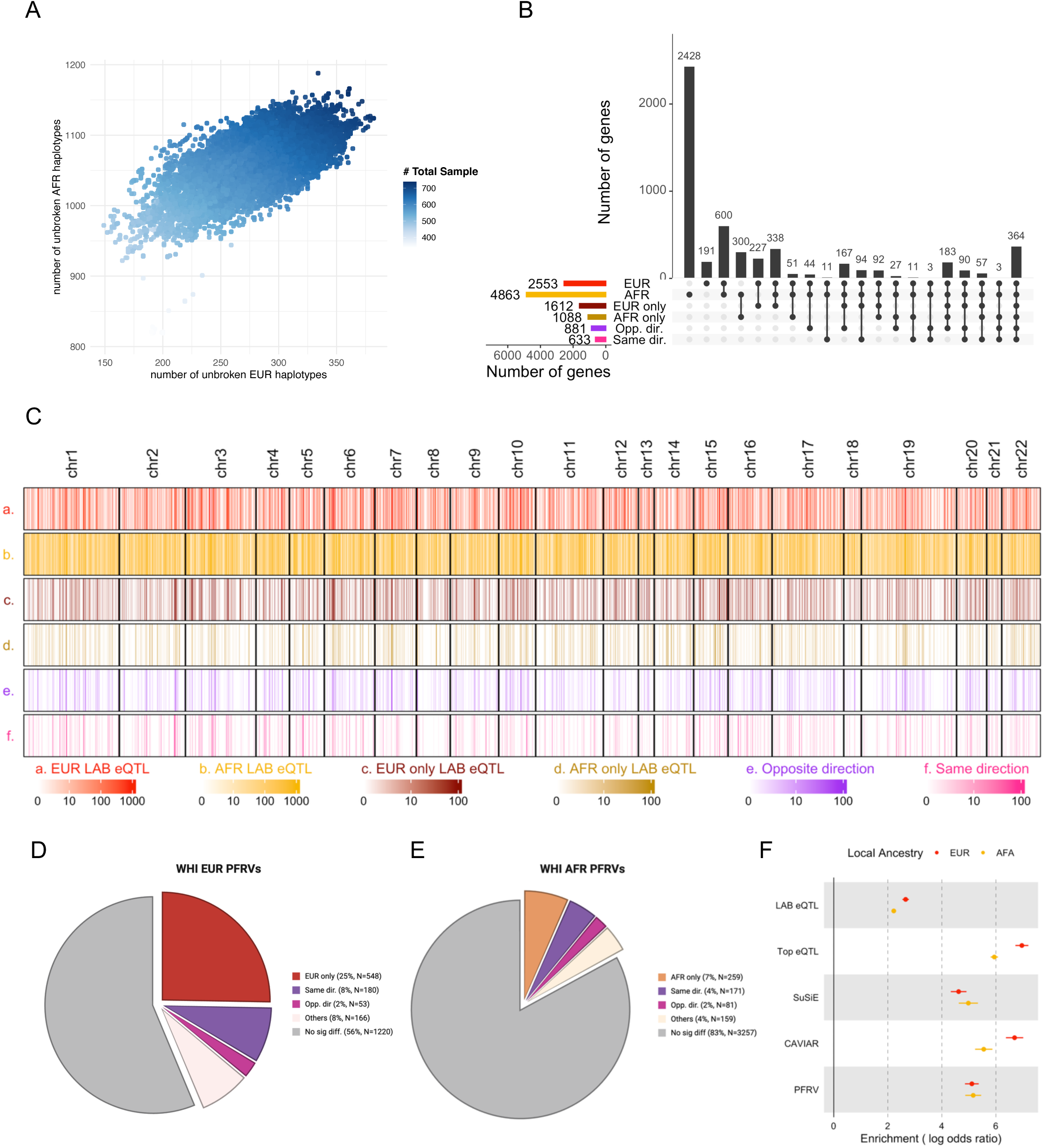
Local ancestry distribution, the identified LAB eQTL and their replication in African American from WHI. A) Estimated global ancestry proportions of WHI participants (autosomes) derived using ADMIXTURE. Each vertical bar represents a single individual, and the four color-coded segments represent the three ancestry fractions. B) Scatter plot of number of unbroken EUR (x-axis) and AFR (y-axis) haplotypes for each gene, the grade of color represents the number of subjects with both unbroken haplotypes. D) Upset plot showing the number of genes with AFR and/or EUR LAB eQTLs as well as different types of local ancestry-differentiated LAB eQTLs in WHI. A single gene may contain multiple types of local ancestry differentiated LAB eQTLs, referring to distinct loci. E) Forest plot shows the enrichment of identified WHI LAB eQTLs in replication as well as the top and fine-mapped LAB eQTLs. The color indicates the source of local ancestry background. The enrichment is estimated with TORUS, and the bar represents the 95% confidence interval of estimation.

### Fine mapping in WHI

We applied the fine-mapping approach described above to the WHI results. The DAP-G method detected no signals in this cohort at the default posterior probability threshold of 0.8, likely due to a combination of method-specific characteristics, the relatively low frequency of EUR alleles in the dataset, and AFR-specific LD structure. We identified 2,167 PFRVs for 955 genes from EUR LAB eQTLs and 3,927 PFRVs for 2,353 genes from AFR LAB eQTLs. Among these variants, 1,573 (26.8%, 1,573/5,865) had had ancestry-specific differences in effect, defined as significantly different effect sizes (FDR < 0.05) across 883 genes, including three patterns, 1) functional in one ancestry with no evidence of function in the other (548 EUR-only and 259 AFR-only PFRVs), 2) functional in both ancestries with the same direction of effect but different effect sizes (309), and functional in both ancestries with opposite directions of effect (132) (**Figures 4D-E, S7 and Table S5)**. In total, 32.7% (883/2,702) of the genes evaluated had at least one PFRV with an ancestry-specific effect.

### Replication of WHI LAB eQTLs

Our replication set for WHI included 346 unrelated participants. Of the 535,767 EUR LAB eQTLs in the discovery stage, 222,745 (41.6%) were validated in the replication set, remaining significant after FDR correction. Similarly, for the AFR haplotypes, 422,081 (48.5%) of 871,129 LAB eQTL were validated, following FDR correction. PFRVs were also highly enriched in the replication set (**Figure 4F**).

### Genetic correlation analyses

We observed increasing genetic correlation of EUR LAB eQTLs with GTEx with increasing heritability of gene expression in the BHRC, reaching 0.91 (IQR: 0.75-0.97) for genes with h^2^ above 0.4 (**Figure 5A** and **Table S6**). By contrast genetic correlation of EUR LAB eQTLs with NIA LAB eQTLs is low (0.64, IQR: 0.41-0.82) even for genes with high heritability (h^2^>0.4). Similarly, genetic correlation of highly heritable genes is markedly higher (0.79, IQR: 0.61-0.94 vs 0.67, IQR: 0.46-0.83) between WHI EUR LAB eQTLs with GTEx compared to WHI EUR LAB eQTLs with WHI AFR eQTLs (**Table S7**). These findings suggest that in both populations we observe similar genetic architectures of EUR eQTLs compared to GTEx, a predominantly European ancestry dataset, and lower genetic correlation within a single population between the two admixed ancestries.

**Figure 5.**
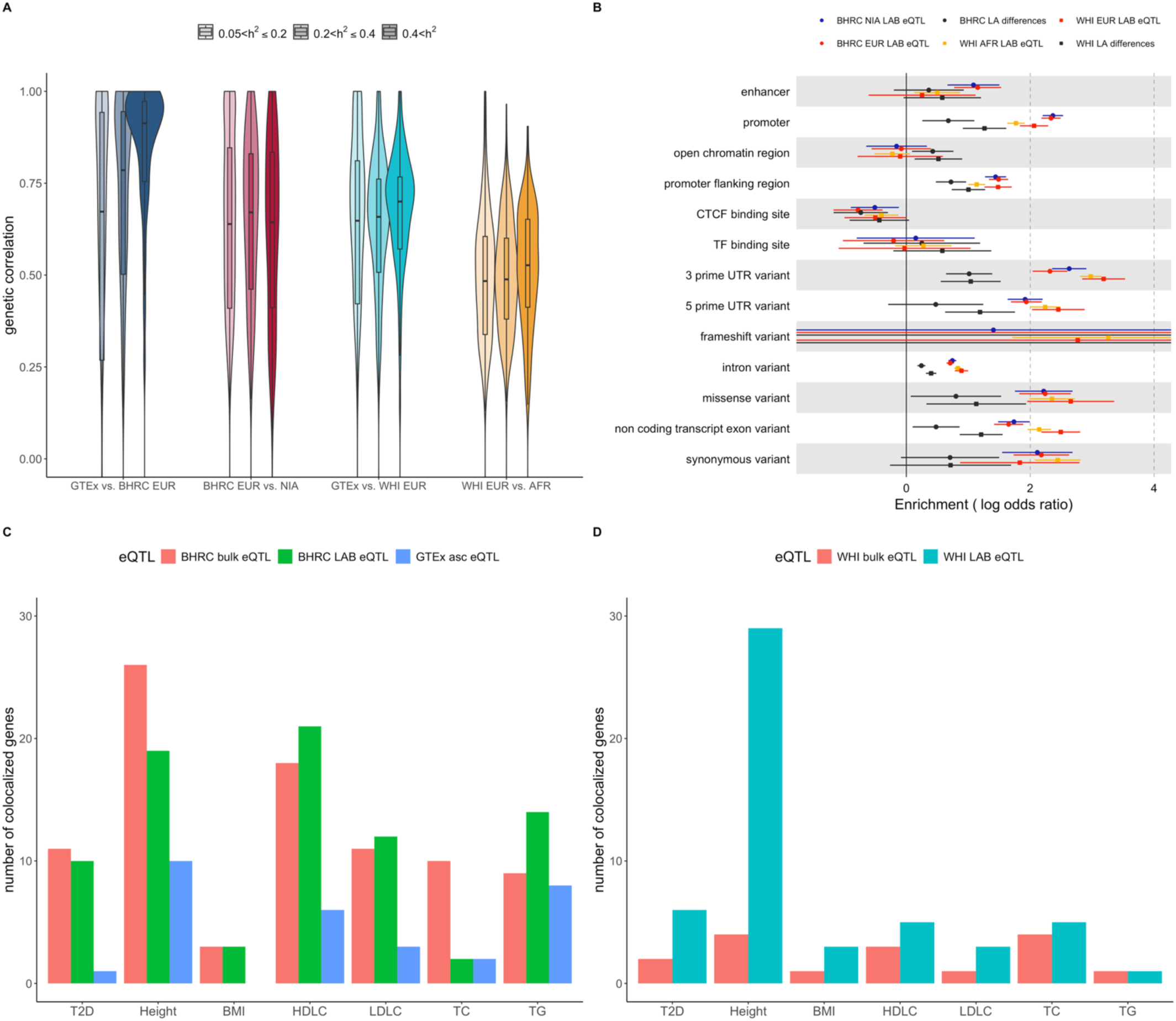
Genetic correlation between local ancestry-based expression quantitative trait loci (LAB eQTLs), functional annotation for LAB eQTLs and coloc analyses for ancestry-matched GWAS with LAB and bulk eQTLs. A) The Violin plot shows the distribution of gene-level genetic correlation of different comparisons, Border Health Research Cohort (BHRC) European ancestry (EUR) vs. GTEx (dark blue) BHRC EUR vs. Native/Indigenous American ancestry (NIA) (dark red), Women Health Initiative (WHI) European ancestry (EUR) vs. GTEx (blue), and WHI EUR vs. African ancestry (AFR) grouped by different levels of heritability of gene expression, estimated by GNOVA (0.05≤h^2^<0.2, 0.2≤ h^2^<0.4, and h^2^≥0.4). An trend of increased genetic correlation with increased heritability was observed in the comparison between BHRC/WHI EUR vs. GTEx; this trend was not observed in the comparison between BHRC EUR vs. NIA or WHI EUR vs. AFR. B) The forest plot shows the enrichment score of LAB eQTLs for two local ancestry backgrounds (red for EUR, blue for NIA, and yellow for AFR) and differences between them (black, measured by, which is equal to in Eq. 1) for each functional annotation from Ensembl Regulation. The bar indicates the 95% confidence interval. Both EUR and NIA LAB eQTLs are enriched in enhancer, promoter, and other functional regions. In contrast, only those eQTLs with more significant differences by ancestry are enriched in open chromatin regions. C) The bar chart shows the number of identified colocalized genes of Hispanic/Latino GWAS with different eQTL mapping approaches and source populations, including LAB and bulk eQTLs from BHRC, and ASC eQTLs from GTEx. Every studied trait shows a greater number of colocalized genes with BHRC LAB eQTLs when compared to ASC eQTLs from GTEx. D) The bar chart shows the number of identified colocalized genes of African American GWAS with different eQTL mapping approaches, including LAB and bulk eQTLs from WHI.

### Genome region annotation

To classify the function of LAB eQTLs, we leveraged the Ensembl Regulatory Build(*14*) to identify regulatory elements. Among the three million LAB eQTLs we identified in BHRC, both EUR and NIA LAB eQTLs were enriched in promoter and enhancer regions and depleted in CTCF binding sites. We also observed similar enrichment results for AFR and EUR LAB eQTLs in WHI. In contrast, in both the BHRC and WHI, the LAB eQTLs with significant differences in ancestry-specific effect sizes were found to be significantly enriched in open chromatin regions, while other LAB eQTLs were not (**Figure 5B**).

### Colocalization analyses

Finally, we tested the hypothesis that our LAB eQTLs provide greater understanding of the molecular basis of GWAS associations in admixed populations using colocalization analyses of eQTLs with ancestry-specific effect size differences for type 2 diabetes (T2D), four lipid traits (HDL, LDL, triglycerides, total cholesterol), height, and body mass index (BMI).(*15–17*) Among Hispanic/Latino GWAS, for all traits except total cholesterol, LAB eQTLs colocalized to more regions than the GTEx-derived map (**Figures 5C** and **S8**), with 10, 19 and three colocalized regions for T2D, height, and BMI, compared to 1, 10, and 0 regions, respectively, for GTEx allele-specific count eQTLs (ascQTLs). Across the four lipid traits, our ancestry-specific eQTL analyses identified a total of 58 colocalized regions, while the GTEx ascQTL analyses only identified 19 colocalized regions. Similarly, among African American GWAS, LAB eQTLs colocalized to a higher number of loci compared to traditional eQTL mapping approaches, e.g., 29 and 6 colocalized genes were identified for height and T2D with LAB eQTLs compared to 4 and 2 with traditional eQTLs (**Figures 5D** and **S8**). Similarly, our BHRC LAB eQTL map outperformed traditional eQTL mapping based on the same transcriptomic data for some traits; for example, nine regions colocalized with the BHRC traditional eQTL map for triglyceride GWAS compared to 14 with LAB QTLs. However, for total cholesterol, body height, and T2D, LAB eQTLs identified fewer colocalized genes compared to traditional eQTLs, albeit with an average of 17.17-32.4% reduction in sample size and only using only 5.88-9.96% of sequencing reads for *ancQTL*.

## Discussion

In this study, we introduce a new method for eQTL mapping that explicitly accounts for local ancestry in admixed populations, *ancQTL*. We generated the first-ever profile of a local ancestry-specific regulatory landscape in two studies with predominantly two-way admixed populations, a Hispanic/Latino cohort, BHRC, primarily comprising NIA and EUR ancestry, and an African American cohort, WHI, comprising predominantly AFR and EUR ancestry. This regulatory landscape is characterized by pervasive variant effect differences by underlying ancestry, with 45.6% and 52.3% of eGenes (i.e., genes with an LAB eQTL in these analyses) having differential eQTL effects by local ancestry, in WHI and BHRC respectively.

These extensive ancestry-specific effect differences alone do not imply that that there are ancestry-specific differences in the regulatory function of the underlying causal variants mechanistically influencing gene expression. These results could instead be explained solely by differences in causal allele population frequencies and population LD patterns. To test this hypothesis, we applied three state-of-the-art fine mapping tools to identify the union set of PFRVs that were likely to mechanistically influence gene expression. While the proportion of eGenes with PFRVs that exhibit differential effects by local ancestry was reduced relative to the proportion observed from overall LAB eQTLs, PFRVs with ancestry differences were nonetheless observed throughout the whole genome (**Figures 3D-E**), suggesting that ancestry-specific regulatory function is present in as many as a third of genes (32.7% and 41.9% in WHI and BHRC, respectively).

These heterogeneous regulatory effects by local ancestry provide a mechanistic explanation for inconsistent and even incorrect interpretation of GWAS,(*18*) and may improve the informativeness of transcriptome-wide association studies(*19*) and polygenic risk scores across populations.(*20, 21*) For example, LAB eQTLs with opposing effect direction on expression, and thus disease, will elude discovery in traditional logistic or linear GWAS frameworks of admixed populations. We expect this class of variants may disproportionately contribute to disease risk(*22, 23*) and indeed, our study finds a notable increase of colocalization of LAB eQTLs with GWAS signals relative to traditional GTEx eQTLs (**Figure 5A**), improving functional interpretation of genetic risk for several complex traits including T2D, blood lipids, body height, and BMI.

Several possible molecular mechanisms may underlie the observed differential effects by ancestry and their colocalization with disease loci. The first possible explanation is that local ancestry effects may describe the aggregate effects of the underlying haplotype tagged by the allele of interest, with variable haplotypes in different ancestral backgrounds causing different effect estimates at the same variant. Secondly, transcriptional regulation can be gene-specific and depends on a variety of both local and distant factors.(*24*) In our functional annotation, we observed that eQTLs with distinct effects by ancestral background were enriched in open chromatin regions in both BHRC and WHI, suggesting these key components of the epigenetic landscape may be involved in ancestry-specific gene regulation. In line with these findings, a recent study of local ancestry underlying differential chromatin accessibility and single nuclei transcription levels found that 40% of significantly differentially expressed genes showed significant changes in chromatin accessibility by local ancestry.(*25–27*)

Prior investigations of complex traits in African-European admixed individuals observed high correlation of genetic variants with traits across local ancestries across the genome; this estimated correlation was much higher than correlations across continental ancestries.(*28, 29*) Our results are consistent with these findings. For example, we see a genome-wide correlation of EUR LAB eQTLs with GTEx-derived eQTLs of 0.733 and in NIA of 0.643. Nevertheless, our data suggest that differential LAB eQTL effects are common, even after rigorous fine mapping. While these results may initially seem inconsistent, they are likely explained by the fact that many of the differential LAB eQTLs and PFRVs we identified had consistent direction of effect in both ancestries (**Figure S5**). However, gene expression measures are much closer to the mechanism of gene action than complex disease, and often exhibit larger effect size, increasing our power to detect effect differences. Additionally, RNA-seq data reflects DNA sequence information, allowing us to directly measure the regulation effect specific to the local ancestry underlying a single copy of the gene, which is not possible for analyses of common complex disease. Our approach is also robust to many of the sources of bias identified as potential causes of inflated effect estimates; namely, our sample sizes are roughly balanced in terms of local ancestry (**Figures 1** and **4**) and omics-based cis-QTLs are not highly polygenic.(*28*) Additionally, studies of ancestry effects on gene regulation are often confounded by non-genetic influences. Since we estimate ancestry-specific effects in the same group of individuals, often capturing differences in expression by ancestry within individuals, our results are subject to reduced bias from population-level differences in environment, such as lifestyle, diet, and socioeconomic status. Therefore, the regulatory effect landscape captured by LAB eQTLs better reflects genetic effects influencing cellular functions. Together, these factors may contribute to increased power to detect ancestry differences relative to prior research.(*7, 28, 30*)

Despite the advantages of ancQTL for detecting accurate LAB eQTL effects, many genes contain ancestry-specific breakpoints and many RNA sequencing reads cannot be mapped to specific haplotypes due to lack of genetic variation within the read; these factors diminish statistical power. Here, we only used information from samples in which the cis-region haplotype was from a single ancestry population (unbroken local ancestry, illustrated in **Figure S1A**), which caused an average loss of 32.4% of samples in BHRC and 17.17% in WHI, with sample exclusions varying for each gene (**Figure S1B**). In addition, only sequencing reads with one or more heterozygous sites could be used to distinguish the source haplotype of reads, and as a result, only 9.96% of assigned sequencing reads were included in our BHRC analysis and 5.88% of assigned reads were included in our WHI analysis. Despite these limitations, we still observed strong genetic correlation of EUR LAB eQTLs (from both BHRC and WHI) with GTEx ascQTLs. Nevertheless, a large proportion of identified LAB eQTLs replicated in independent sets from each cohort, demonstrating that, despite reduced power, many of the associations identified by ancQTL are reproducible. Another limitation of our approach is that it models the effects of only two local ancestries in the populations we studied. Extending this framework to incorporate local ancestry effects in populations with three or more ancestral backgrounds and to more fine-grained admixture within continental populations are important directions for future work.

In conclusion, we demonstrated that heterogeneous regulatory effects by local ancestry context are widespread and contribute to inconsistencies and reduced power for interpretation of GWAS findings across admixed populations. We further found that LAB eQTLs improve colocalization and aid in the biological interpretation of genetic effects for a range of complex traits. These findings emphasize the importance of accurate ancestry-matching of eQTL data for interpreting GWAS signals and the need for more and larger transcriptome datasets from diverse populations. For the complex traits we considered, understanding why transcript abundance varies on distinct admixed haplotypes will be instrumental for understanding the etiology of disease disparities, identifying potential prevention measures and therapeutics, and ensuring that all populations benefit from advances in precision prevention and treatment. Indeed, improved understanding of the functional role of regulatory variation in admixed populations has implications for all modern populations. We anticipate that LAB eQTL approaches, such as *ancQTL*, will become increasingly important as global populations become more admixed.(*31, 32*) Our findings underscore the necessity of incorporating local genomic context in genetic studies, with implications for refining models of gene regulation, advancing precision medicine, and addressing health disparities.

## Data Availability

All data produced in the present study are available upon reasonable request to the authors

## Acknowledgements

The authors would like to thank the BHRC team, particularly Rocío Uribe, BSIE, and her team, who recruited and interviewed the participants, Marcela Morris, BS, for laboratory support, the data management team, and Norma Pérez-Olazarán, BBA, and Christina Villarreal, BA, for administrative support; Valley Baptist Medical Center, Brownsville, Texas, for providing us space for our Center for Clinical and Translational Science Clinical Research Unit; and the community of Brownsville and the participants who so willingly participated in this study in their city. E.R.G. is grateful to the President and Fellows of Clare Hall, University of Cambridge for fellowship support. Molecular data for the Trans-Omics in Precision Medicine (TOPMed) program was supported by the National Heart, Lung and Blood Institute (NHLBI). Whole genome sequencing for “NHLBI TOPMed: Women’s Health Initiative (WHI)” (phs001237.v3.p1) was performed at the Broad Institute of MIT and Harvard (3U54HG003067-13S1). RNASeq for “NHLBI TOPMed: Women’s Health Initiative (WHI)” (phs001237.v3.p1) was performed at Broad Institute Genomics Platform. Core support including centralized genomic read mapping and genotype calling, along with variant quality metrics and filtering were provided by the TOPMed Informatics Research Center (3R01HL-117626-02S1; contract HHSN268201800002I). Core support including phenotype harmonization, data management, sample-identity QC, and general program coordination were provided by the TOPMed Data Coordinating Center (R01HL-120393; U01HL-120393; contract HHSN268201800001I). We gratefully acknowledge the studies and participants who provided biological samples and data for TOPMed.

## Ethics

The BHRC portion of this study was approved by the Committee for the Protection of Human Subjects of the University of Texas Health Science Center, Houston TX. The WHI portion of this study was approved by the Institutional Review Board at the Fred Hutchinson Cancer Center, Seattle WA.

---

Figure S1. Illustration of qualified and disqualified haplotypes and their distributions. A) To accurately capture the effect of LAB eQTL, we excluded haplotypes without continuous local genetic ancestry of either Native/Indigenous American (NIA) or European (EUR) spanning the cis-regulation region (within 1Mb of transcription start site) of the target gene. B) Histogram showing the number of genes by the proportion of haplotypes without continuous local ancestry among 645 subjects (1,290 haplotypes) in our discovery set. C) The scatterplot presents the average NIA global ancestry proportion for subjects with disqualified haplotypes and the proportion of haplotypes without continuous local ancestry.

Figure S2. Volcano and QQ plots for the global ancestry effect on gene expression. The volcano plot shows the effect size (beta) of global ancestry, representing with first the PC, on the gene expression with the adjustment of sex, age, RNA-seq batch, and 60 PEER factors. Red dots indicate genes with false discovery rate (FDR)<0.05.

Figure S3. A. Exemplar scenarios comparing power to detect effects of variants with differences in local ancestry-based expression quantitative trait locus (LAB eQTL) effect sizes by local ancestry on disease risk. B. The Mendelian randomization (MR) model used to assess the reduction in power to detect causal effects when local ancestry-based effect differences are ignored. We find power to detect disease effects is reduced when eQTL effect sizes differ by local ancestry.

Figure S4. A) Power to detect differences in local ancestry based eQTL effects (alpha<0.05) for sample sizes ranging from 100 to approximately our sample size, 650, and diverse transcript abundance (total aligned sequencing reads ranging from 75-1500). The row and column represent the difference of effect in the two ancestry backgrounds, with fold change ranging from 1.01 to 1.5. For each scenario, we conducted 200 simulations to obtain the power. B) Type I error was evaluated under the scenario in which FC1 was equal to FC2.

Figure S5. A) The QQ plot of LAB eQTL p-values for EUR (blue) and NIA (yellow) haplotypes as well as the p-values of the G_A_*G_LA_ interaction effect (green), and the p-values for asc eQTL (orange), showing expected levels of inflation, in line with Liang et al.(*22*) B) The Venn diagram of identified LAB eQTLs and asc eQTLs, and their overlap with variants with significant G_A_*G_LA_ interaction effects.

Figure S6. A) Venn diagram of the fine-mapped LAB eQTLs for EUR (left) and NIA (right) in BHRC. The two plots indicate the numbers of identified causal LAB eQTLs, defined with posterior probability >0.8, with three different tools, including SuSiE, DAP-G, and CAVAIR, and their overlap B) The heatmap summarizes the number of identified PFRVs for each gene and different types of local ancestry differentiated LAB eQTLs in BHRC. C) Upset plot showing the number of genes with EUR and/or NIA PFRVs as well as different types of local ancestry-differentiated LAB eQTLs in BHRC. A single gene may contain multiple types of local ancestry differentiated LAB eQTLs, referring to distinct loci.

Figure S7. A) Venn diagram of the fine-mapped LAB eQTLs for EUR (left) and AFR (right) in WHI. The two plots indicate the numbers of identified causal LAB eQTLs, defined with posterior probability >0.8, with SUSiE and CAVIAR and their overlap. The DAP-G method detected no signals in this cohort at the default posterior probability threshold of 0.8, likely due to a combination of method-specific characteristics, the relatively low frequency of EUR haplotypes in the dataset, and AFR-specific LD structure. B) The heatmap summarizes the number of identified PFRVs for each gene and different types of local ancestry differentiated LAB eQTLs in WHI. C) Upset plot showing the number of genes with EUR and/or NIA PFRVs as well as different types of local ancestry-differentiated LAB eQTLs in WHI. A single gene may contain multiple types of local ancestry differentiated LAB eQTLs, referring to distinct loci.

Figure S8. Coloc analysis for Hispanic/Latino GWAS with LAB eQTLs from BHRC and asc eQTLs from GTEx. The Miami plots show the identified colocalized genes with BHRC LAB eQTLs and GTEx ASC eQTLs from H/L type 2 diabetes (A), height (B) BMI (C), HDL cholesterol (HDLC) (D), LDL cholesterol (LDLC) (E), triglycerides (TG) (F) and total cholesterol (TC) (G).

